# Analysis of phage therapy failure in a patient with a *Pseudomonas aeruginosa* prosthetic vascular graft infection

**DOI:** 10.1101/2023.03.19.23287399

**Authors:** Lucia Blasco, Inmaculada López-Hernández, Miguel Rodríguez-Fernández, Javier Pérez-Florido, Carlos S. Casimiro-Soriguer, Sarah Djebara, Maya Merabishvili, Jean-Paul Pirnay, Jesús Rodríguez-Baño, María Tomás, Luis Eduardo López Cortés

## Abstract

Clinical case of a patient with a *Pseudomonas aeruginosa* multidrug-resistant prosthetic vascular graft infection which was treated with a cocktail of phages (PT07, 14/01 and PNM) in combination with ceftazidime-avibactam (CAZ/AVI). After the application of the phage treatment and in absence of antimicrobial therapy, a new *P. aeruginosa* bloodstream infection (BSI) with a septic residual limb metastasis occurred, now involving a wild-type strain being susceptible to ß-lactams and quinolones. Clinical strains were analyzed by microbiology and whole genome sequencing techniques. In relation with phage administration, the clinical isolates of *P. aeruginosa* before phage therapy (HE2011471) and post phage therapy (HE2105886) showed a clonal relationship but with important genomic changes which could be involved in the resistance to this therapy. Finally, phenotypic studies showed a decreased in Minimum Inhibitory Concentration (MIC) to ß-lactams and quinolones as well as an increase of the biofilm production and phage resistant mutants in the clinical isolate of *P. aeruginosa* post phage therapy.

**Importance:** Phage therapy is a promising new treatment against infections produced by multi-drug resistant pathogens. For that, it would be necessary to know more about the clinical response and host-phage interactions by massive sequencing techniques to improve phage therapy application. In this work, we analyzed the clinical, microbiological and molecular features of the *P. aeruginosa* isolates in prosthetic vascular graft infection after the phages administration failure against this infection. This knowledge could allow to develop strategies of improvement of the use of phage therapy as treatment of multiple clinical infections.

---

We present the case of a in their 50s man who in July 2020 developed a surgical infection caused by ceftazidime and piperacillin-tazobactam resistant *Pseudomonas aeruginosa*. In August 2020, an axillo-bifemoral bypass was performed. Shortly afterwards, the patient developed a bloodstream infection (BSI) caused by *P. aeruginosa* with the same antibiotic resistance pattern, originating from a sacral ulcer, which was treated with meropenem (1g, q8h for 17 days). The patient suffered up to five recurrences of *P. aeruginosa* BSI between October and December 2020, while the sacral ulcer developed favourably. After the second relapse, 18F-fluorodeoxyglucose (18F-FDG) PET/CT was performed, showing pathological radioactive-labelled glucose uptake along the preclavicular graft region (SUVmax value, 5.29) and suggesting prosthetic vascular graft infection (PVGI). Nevertheless, surgical management was rejected. The patient subsequently received different targeted antibiotherapies for two to six weeks (Figure 1). Two new *P. aeruginosa* BSI relapses occurred between March and May 2021. Ceftolozane-tazobactam was not used (not available), and off-label tebipenem use was denied; treatment was therefore as shown in Figure 1. Simultaneously, therapeutic phages were obtained from the Queen Astrid Military Hospital (Brussels, Belgium) in view of a possible compassionate use phage therapy. Phage susceptibility of the clinical isolate was determined in the A Coruña University Hospital, and three *P. aeruginosa* phages (PT07, 14/01 and PNM) were selected [1, 2]. In July 2021, 70 ml of the bacteriophage cocktail, consisting of the three phages, each at a concentration of 10^7^ plaque forming units (PFU)/mL, was administered intravenously, once a day in a 6-hour infusion for three days in an inpatient regimen. No adverse events were observed. Phage therapy was followed by four days of outpatient parenteral antimicrobial therapy (OPAT) at the patient’s request. Ceftazidime-avibactam was applied for 8 weeks (6 weeks before and 2 weeks after the phage therapy). In August 2021, in the absence of antimicrobial therapy, a new *P. aeruginosa* BSI with a septic residual limb metastasis occurred, now involving a wild-type strain being susceptible to ß-lactams and quinolones. Finally, upon a multidisciplinary discussion, a proximal vascular prosthesis replacement combined with antibiotherapy in OPAT was performed. Currently, after ten months without treatment, the patient remains asymptomatic.

**Figure 1.**
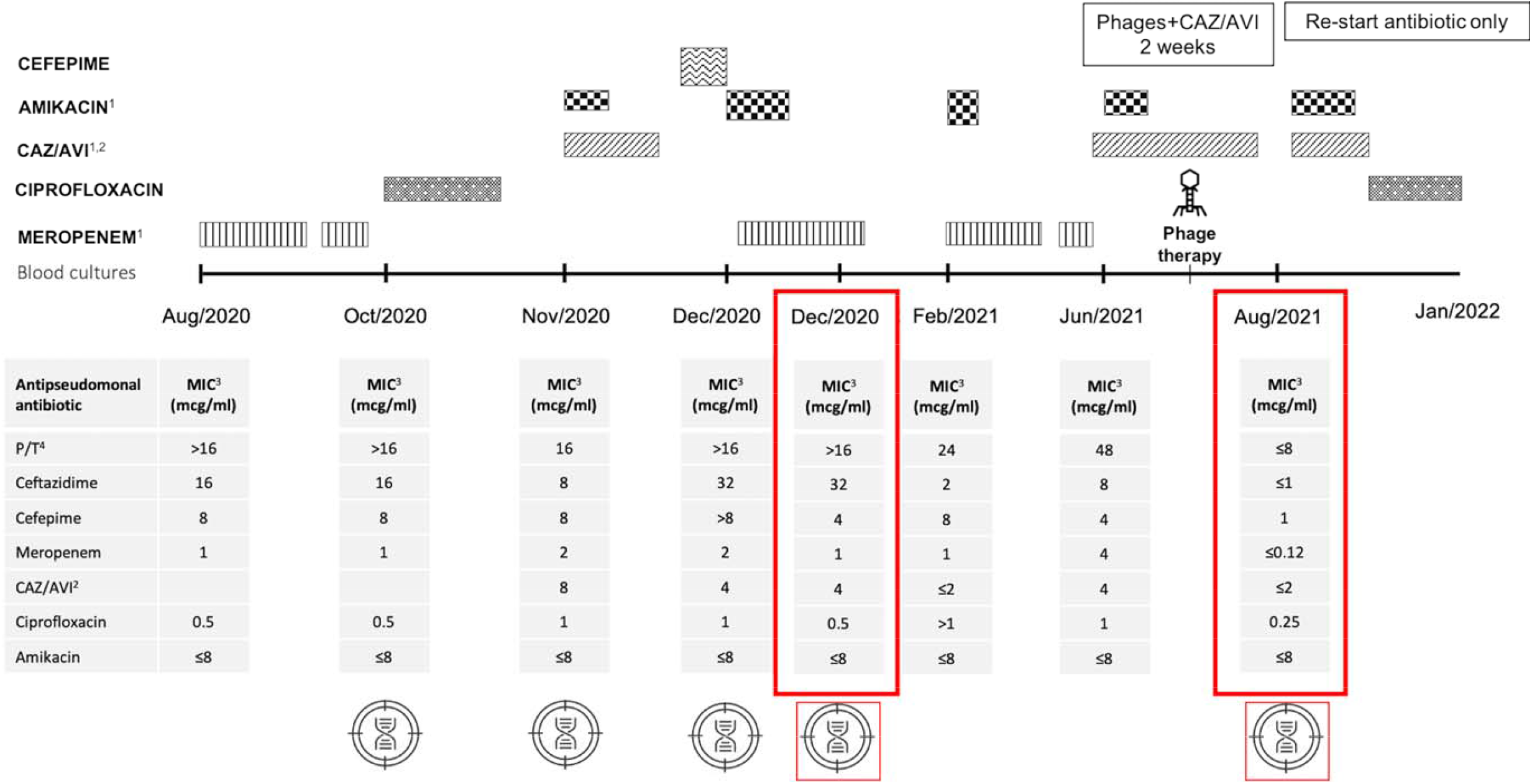
Timeline depicting the different antibiotic and phage therapies, the isolation of *Pseudomonas aeruginosa* strains from blood cultures and their antibiotic susceptibility and genomic analyses. PFGE of these strains in described in the Figure S1. ^1^Completed in outpatient parenteral antibiotic treatment. ^2^CAZ/AVI: Ceftazidime-avibactam. ^3^MIC: Minimum inhibitory concentration. ^4^P/T: Piperacillin-tazobactam. Whole Genome Sequences (WGS) is shown in Black Genome Sequences (three clinical isolates) and Red Genome Sequences, which are bacterial genomes of *Pseudomonas aeruginosa* isolates in relation with phage therapy (two clinical isolates): HE2105886 (after phage treatment) in comparison with HE2011471 (prior to phage administration).

Eight blood culture isolates obtained between November 2020 and August 2021 were characterized by microbiological analysis. These isolates were identified by matrix assisted laser desorption/ionisation - time of flight mass spectrometry (MALDI-TOF MS, Bruker Daltonics) as *P. aeruginosa*. Antimicrobial susceptibility testing was performed by broth microdilution (Microscan, Beckman Coulter) and interpreted using European Committee on Antimicrobial Susceptibility Testing (EUCAST) criteria. Pulsed-field gel electrophoresis (PFGE) revealed that four representative isolates from 2020 and 2021, were clonal (Figure S1). Genomic sequence of five representative isolates showed that these isolates belonged to the high-risk clone ST308 with core genome MLST (MLST)cgST2675. A search for antimicrobial resistance genes in the ResFinder database did not identify any acquired resistance genes such as ß-lactamases. Analysis of 40 chromosomal resistance genes revealed natural polymorphisms in numerous genes, including some changes with unknown effect [3]. The profile of all isolates was identical for all genes studied, except for the *nfxB* gene (a transcriptional repressor that regulates MexCD-OprJ), in which multiple amino acid changes were observed in the last 3 isolates (Table S1, Figure S2). Moreover, comparison of the genomes of isolates HE2011471 (previous to phage administration) and HE2105886 (one month after phage treatment) revealed important mutations (Figure 2). Interestingly, several detected genomic changes could be involved in phage resistance (Table S2, Figure 3). Isolate HE2105886, recovered one month after phage treatment, exhibited a decrease in the Minimum Inhibitory Concentration (MICs) of ß-lactam and quinolone antibiotics (Figure 1), an increased biofilm formation (Figure S3A), and an increased frequency of occurrence of phage resistant mutants (Figure S3B) [4].

**Figure 2.**
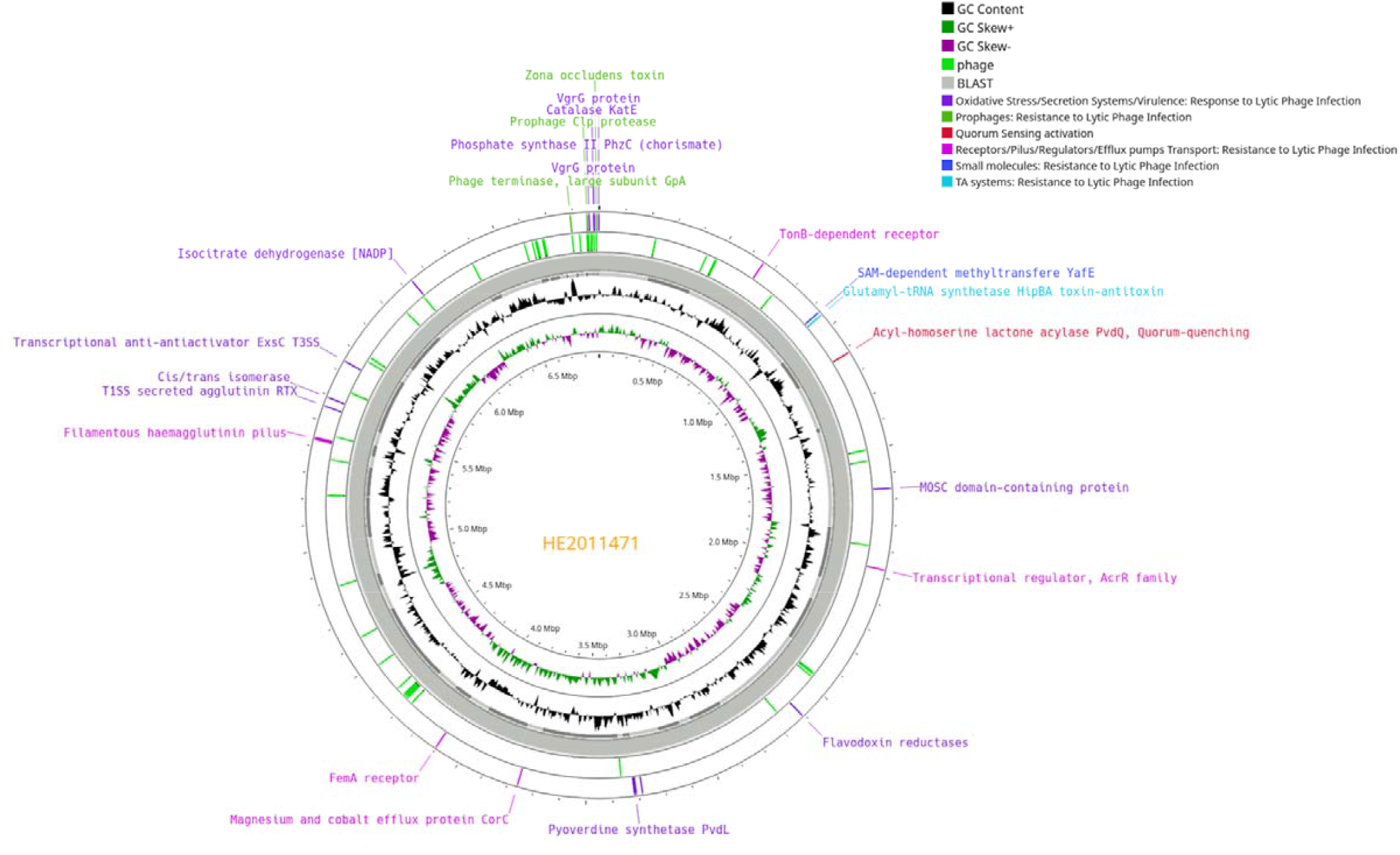
Circular chromosomic view of the bacterial genomes of two *Pseudomonas aeruginosa* isolates: HE2105886 (after phage treatment) in comparison with HE2011471 (prior to phage administration).

**Figure 3.**
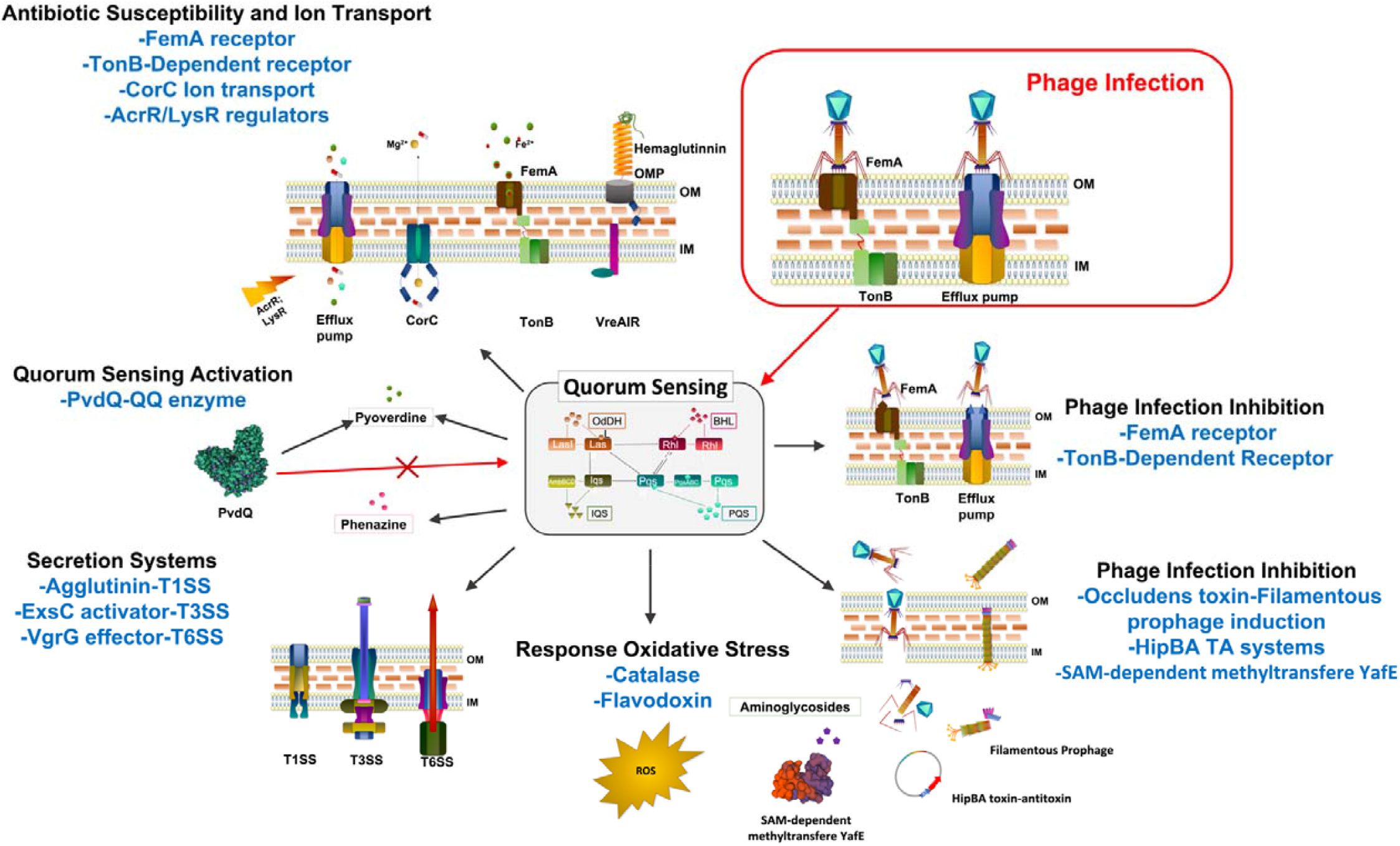
Graphic representation of the proteins coded by the HE2105886 genome with an emphasis of changes associated with mechanisms of resistance and response to phage infection.

## DISCUSSION

This case highlights three key issues. The first relates to the use of combined therapy versus monotherapy as a definitive treatment for *P. aeruginosa* BSI. So far, combination therapy has not been associated with reduced mortality or any advantages in terms of clinical outcome or successful treatment of recurrent/persistent bacteraemia [5]. The second issue concerns prosthetic vascular graft infection (PVGI) diagnosis and its best management. At the early stages of PVGI, a high degree of suspicion is essential. Indium-111-labelled white blood cell scintigraphy plus single-photon emission computed tomography (SPECT/CT) could reduce the false positive rates observed with PET/CT [6]. In the present case, PET/CT was not performed at the early postoperative stage, because of the possibility of a false positive result. Regarding the treatment, graft excision is the preferred surgical approach. However, some patients are considered unacceptable surgical candidates due to underlying comorbidities or technically unfeasible surgery. When this occurs, lifelong suppressive antimicrobial therapy is an option, but is not free of side-effects or the risk of development of antimicrobial resistance. As the patient was initially denied surgical treatment, long-term suppressive treatment with quinolones was administered. However, ciprofloxacin-resistance developed, and surgery was finally deemed key to a favourable outcome. Furthermore, there is no evidence-based recommendation for either PVGI antimicrobial treatment or its optimal duration; minimum intravenous therapy for 6 weeks followed by oral antibiotherapy for up to 6 months has been proposed [7, 8]. ^18^F-FDG PET/CT-guided treatment duration seems feasible and would allow treatment to be tailored to individual patients [9]. The third highlighted issue is related to phage therapy. Phage therapy involves the targeted application of strictly virulent phages that can specifically infect and lyse the targeted pathogenic bacteria they encounter, hereby releasing virion progeny that continues the lytic cycle. A major advantage of phages is the minimal impact on non-target bacteria or body tissues [10]. A recent systematic review suggested that phage therapy is safe and may be effective in different difficult-to-treat infections [11]. Interestingly, a previously-reported case of ß-lactam resistance in an MDR *P. aeruginosa* isolate causing an aortic graft infection was reverted after treatment with a phage that appeared to utilize the outer membrane protein M (OprM) of the MexAB- and MexXY-multidrug efflux pumps, associated with antibiotic resistance, a as receptor, and which was no longer expressed in the selected phage-resistant bacterial strain [12]. In the present case, even though phage therapy did not cure the infection, the posterior infection recurrence was caused by an antibiotic susceptible isolate that belonged to the same lineage as the one that was causing the pre-phage treatment episodes of infection. However, the recurrent isolate was recovered one month after the phage therapy, and it is possible that the resensitization (to ß-lactams and quinolones) could have been due to a spontaneous evolution of the resident strain. Further studies are necessary to elucidate whether the genomic changes found in the bacteria are responsible for the change in phenotype and are related to the phage therapy. One strength of this study is that phage therapy was able to be safely administered in OPAT for greater patient convenience. To our knowledge, this has only been reported once before, in a series of six cases, without phage-related adverse events [13]. The main limitations our study were: first, we did not perform in vitro studies of the effect of the phages in combination with antibiotics, specially ß-lactams; second, we could not study the *P. aeruginosa* clinical isolates during the seven days of application of phage therapy. Finally, it should be noticed that there are no national or local phage bank with characterized phages against *P. aeruginosa* in which specific phages could be selected to provide personalized approach in patients with complex infections caused by these bacteria. In conclusion, in complex *P. aeruginosa* infections the choice of antibiotic therapy and its duration are crucial for minimizing antibiotic pressure and development of resistance. Although we have not demonstrated that phage treatment was effective in this case, studying the molecular mechanisms of resistance to phages and bacteria-phage interactions are key to improving phage therapy in the near future.

## MATERIAL/METHODS

### Microbiology studies

The isolates were identified by MALDI-TOF MS (Bruker Daltonics). Antimicrobial susceptibility testing was performed by broth microdilution with a Microscan system (Beckman Coulter), and the results were interpreted according to the clinical breakpoints defined by the European Committee on Antimicrobial Susceptibility Testing (EUCAST). PFGE analysis was performed using *Spe*I.

### Genomic sequencing and bioinformatic tools

Whole genome sequencing of the five isolates was performed in a HiSeq 2500 sequencing system (Illumina), and sequences were assembled using SPAdes 3.10.1.

Clonality testing of genomes was carried out using MLST Finder (from CGE, available at https://bitbucket.org/genomicepidemiology/mlst/src/master/) and Ridom SeqSphere+ v8.5, and the resistance mechanisms were analyzed using the ResFinder database. SNP calling was performed using snippy v4.6.0 against the NCBI PGAP annotation (https://www.ncbi.nlm.nih.gov/genome/annotation_prok/). The assemblies were visually inspected using Bandage v0.8.1. (https://rrwick.github.io/Bandage/). Further functional annotation genes and prophage elements were confirmed using Blastx (http://blast.ncbi.nlm.nih.gov), Hhmer (http://hmmer.org) and also the HHpred tool (https://toolkit.tuebingen.mpg.de/tools/hhpred), which predict functions through protein structure.

Assembled genomes of *P. aeruginosa* isolates were analyzed using Phaster (PHAge Search Tool Enhanced Release) software (https://phaster.ca/) and SourceFinder (https://cge.food.dtu.dk/services/SourceFinder/).

### Biofilm production

Overnight cultures of *P. aeruginosa* isolates HE2011471 and HE2105886 were diluted 1:100 and used to inoculate 100 μL of LB broth in a 96 multi-well plate. The plate was incubated for 24 h at 37 ºC in darkness. The supernatant was discarded and the wells were washed with PBS. One hundred μL of methanol was then added to each well and discarded after 10 min. When the methanol had completely evaporated, 100 μL of crystal violet (0.1%) was added and discarded after 15 min. Finally, the wells were washed with PBS before the addition of 150 μL of acetic acid (30%), and the absorbance was measured at OD 585 nm.

### Frequency of occurrence of phage resistant mutants

The frequency of occurrence of phage resistant mutants was determined as previously described, with some modifications [14]. Overnight cultures of isolates HE2011471 and HE 2105886 were diluted 1:100 in LB and grown to an OD600nm of 0.6–0.7. An aliquot of 100 μL of the culture containing 10^8^ colony forming units (CFU)/mL was serially diluted, and each dilution was mixed with 100 μL of 10^9^ PFU/mL phage cocktail and then plated by the agar overlay method [15]. The plates were incubated at 37 °C for 18 h and the number of CFUs was counted. The frequency of occurrence of phage resistant mutants and phage resistant mutants was calculated by dividing the number of resistant bacteria by the total number of sensitive bacteria.

## Data Availability

All data produced in the present work are contained in the manuscript

## ACKNOWLEDGEMENTS

We are grateful to Juan Manuel Carmona-Caballero (Hospital Universitario Virgen del Rocío), Elena Fraile (Hospital Universitario Virgen del Rocío), Vicente Merino-Bohorquez (Hospital Universitario Virgen Macarena) and Zaira Palacios-Baena (Hospital Universitario Virgen Macarena) for clinical assistance. We also thank to Carlos Velázquez-Velázquez (Hospital Universitario Virgen Macarena) and Jose Miguel Barquero-Aroca (Hospital Universitario Virgen Macarena) for the surgical assistance. Phage studies were financed by grants PI19/00878 and PI22/00323 awarded to M. Tomás within the State Plan for R+D+I 2013-2016 (National Plan for Scientific Research, Technological Development and Innovation 2008-2011) and co-financed by the ISCIII-Deputy General Directorate for Evaluation and Promotion of Research—European Regional Development Fund “A way of Making Europe” and Instituto de Salud Carlos III FEDER, Spanish Network for the Research in Infectious Diseases (CIBER CB21/13/00012 and CIBER project PMP22/00092), and by the Study Group on Mechanisms of Action and Resistance to Antimicrobials, GEMARA (SEIMC, http://www.seimc.org/). L.E.L.C. and J.R.B. were supported by Plan Nacional de I+D+I 2013-2016 and Instituto de Salud Carlos III, Subdirección General de Redes y Centros de Investigación Cooperativa, Ministerio de Economía, Industria y Competitividad, Spanish Network for Research in Infectious Diseases (RD16/0016/0001)-co-financed by European Development Regional Fund ‘A way to achieve Europe, Operative program Intelligent Growth 2014-2020’.

## TRANSPARENCY DECLARATIONS

The authors declare that they have no conflicts of interest.

## DATA AVAILABILITY

All data generated or analyzed during this study are included in this published article [and its supplementary information files]. Bacterial genomes of *Pseudomonas aeruginosa* isolates HE2105886 (after phage treatment) in comparison with HE2011471 (previous to phages administration) are deposited in the GenBank Bioproject PRJNA938043 (https://www.ncbi.nlm.nih.gov/bioproject/?term=PRJNA938043).

## ETHICAL APPROVAL

Medical Direction of Hospital Universitario Virgen Macarena approved the phage therapy in this patient and after signing the informed consent by the patient by use of experimental medication as compassionate use.

## SUPPLEMENTARY MATERIAL

**Figure S1.**
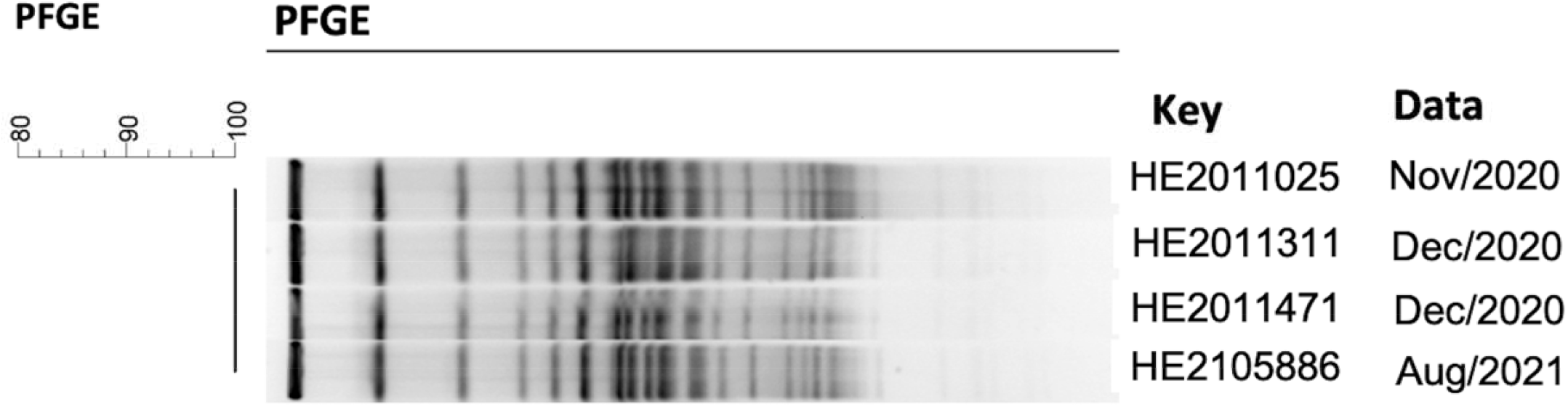
PFGE (*Pulsed Field Gel Electrophoresis*) patterns of four sequential *Pseudomonas aeruginosa isolates*.

**Figure S2.**
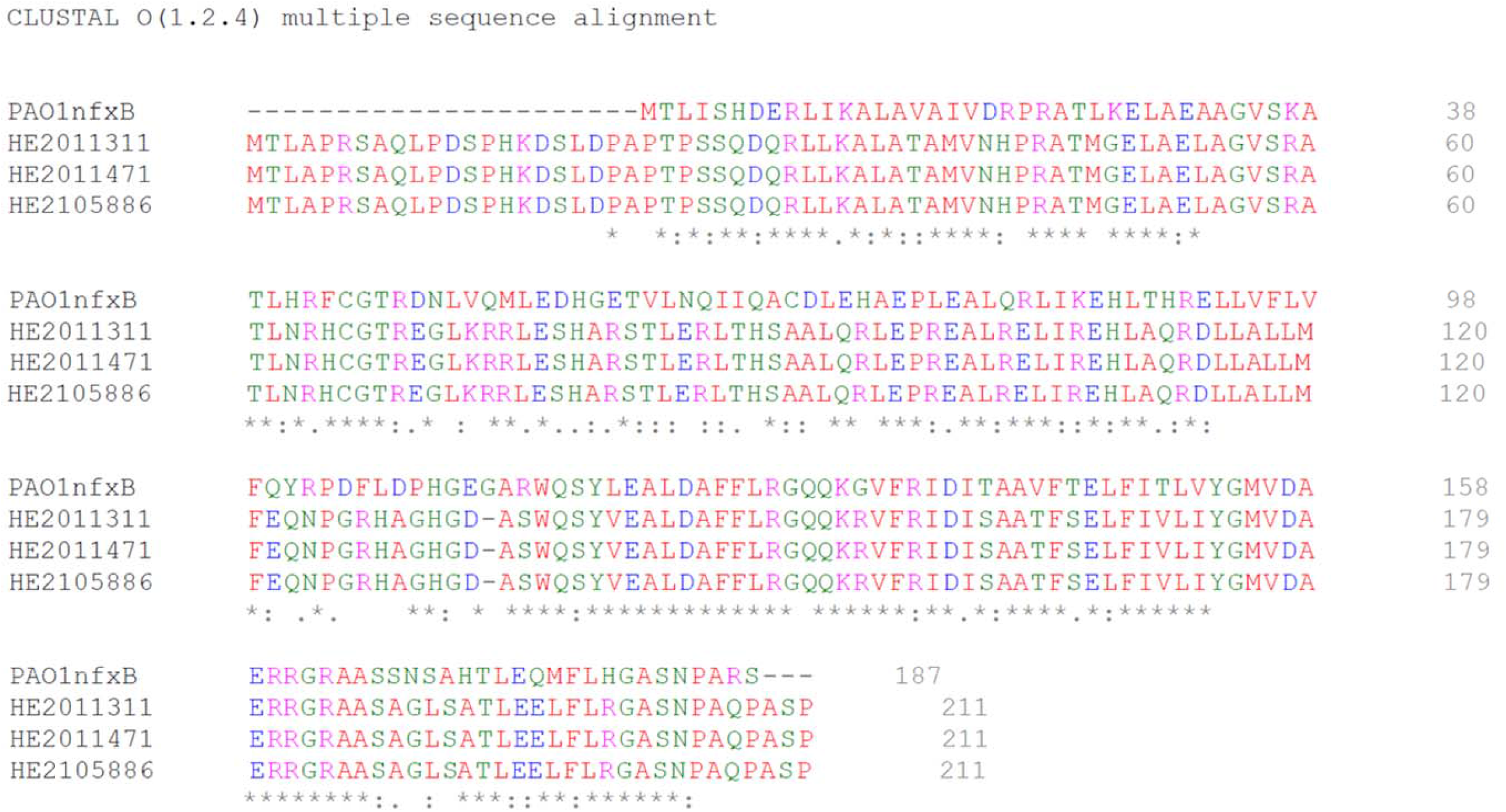
Amino acid changes in NfxB.

**Figure S3.**
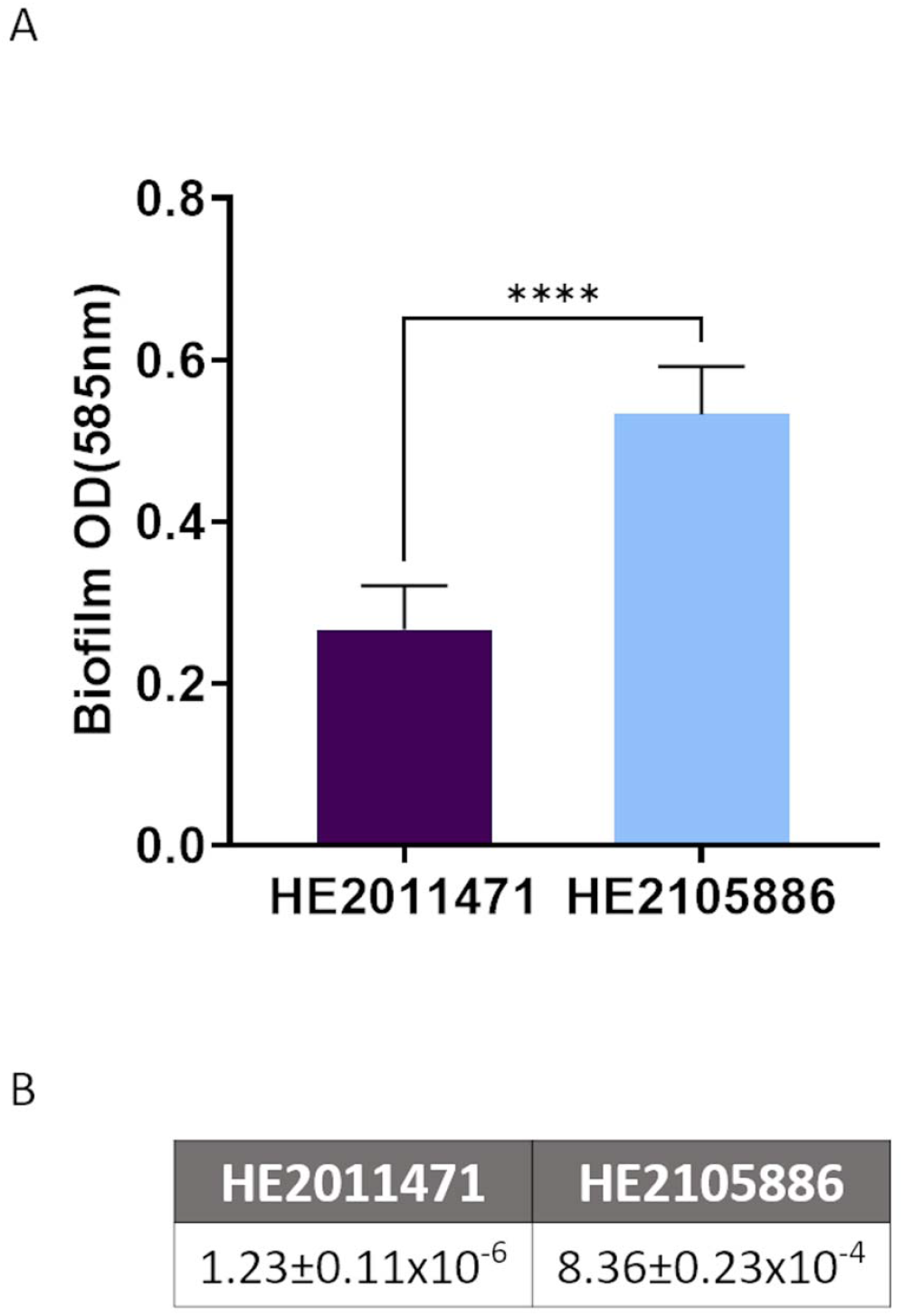
Comparison of the biofilm production (A) and the frequency of occurrence of phage resistant mutants(B) between the *Pseudomonas aeruginosa* isolates HE2011471 (pre phage therapy) and HE2105886 (post phage therapy). **** p value < 0.001 (GraphPad Prism 9.0.0).

**Table S1.**
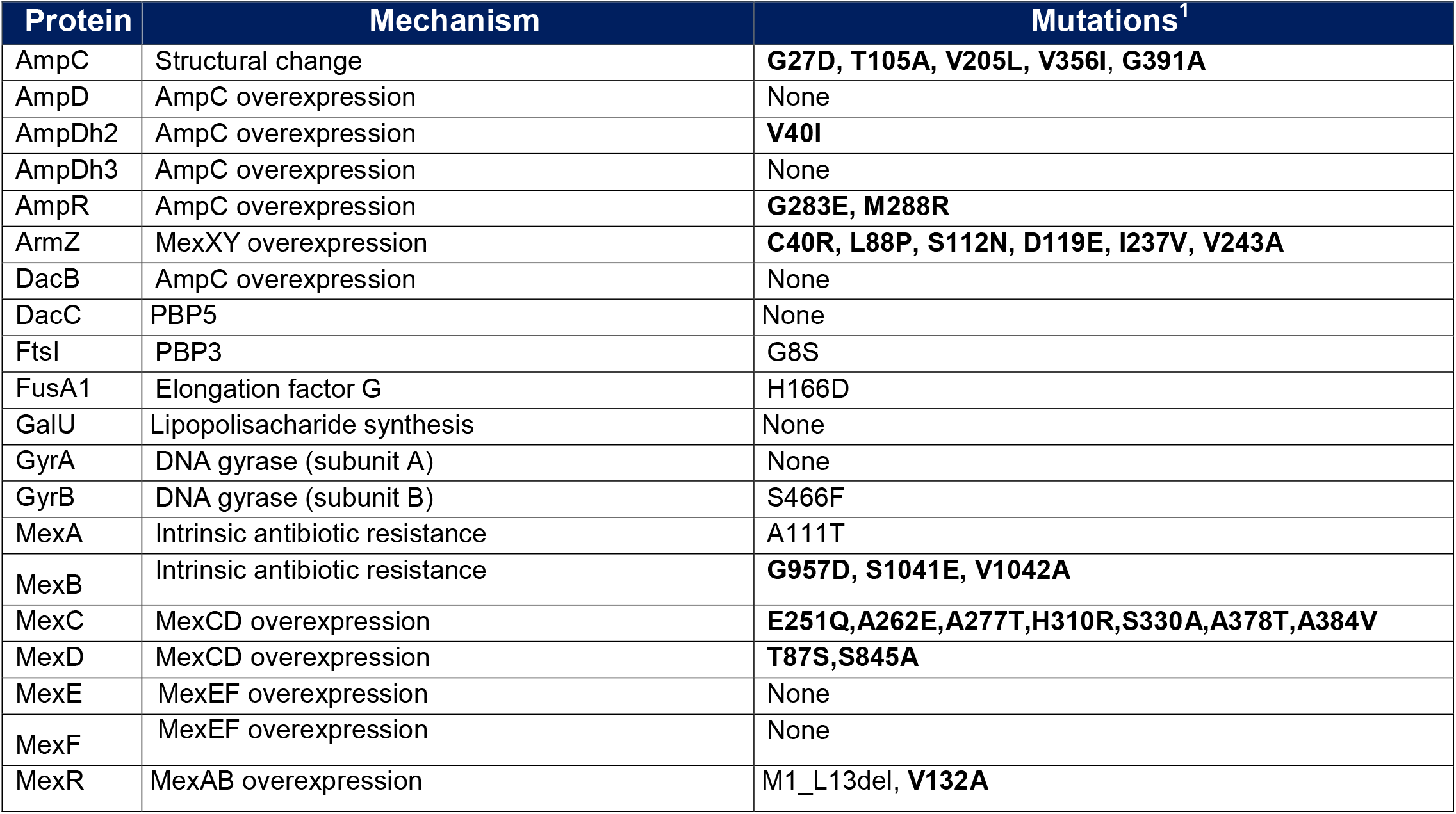
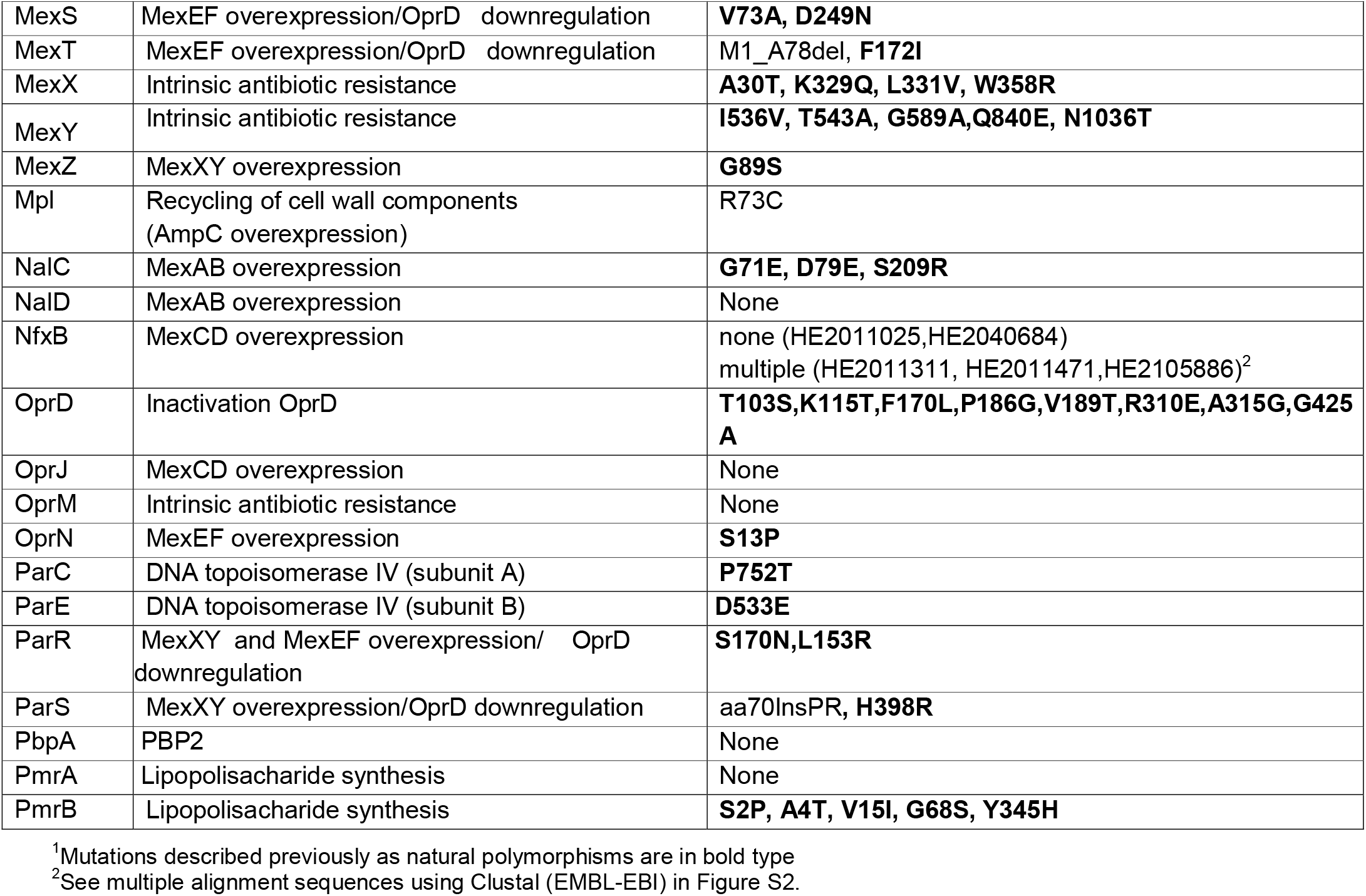
Mutations encountered in proteins known to be related to antimicrobial resistance in *Pseudomonas aeruginosa*.

**Table S2.**
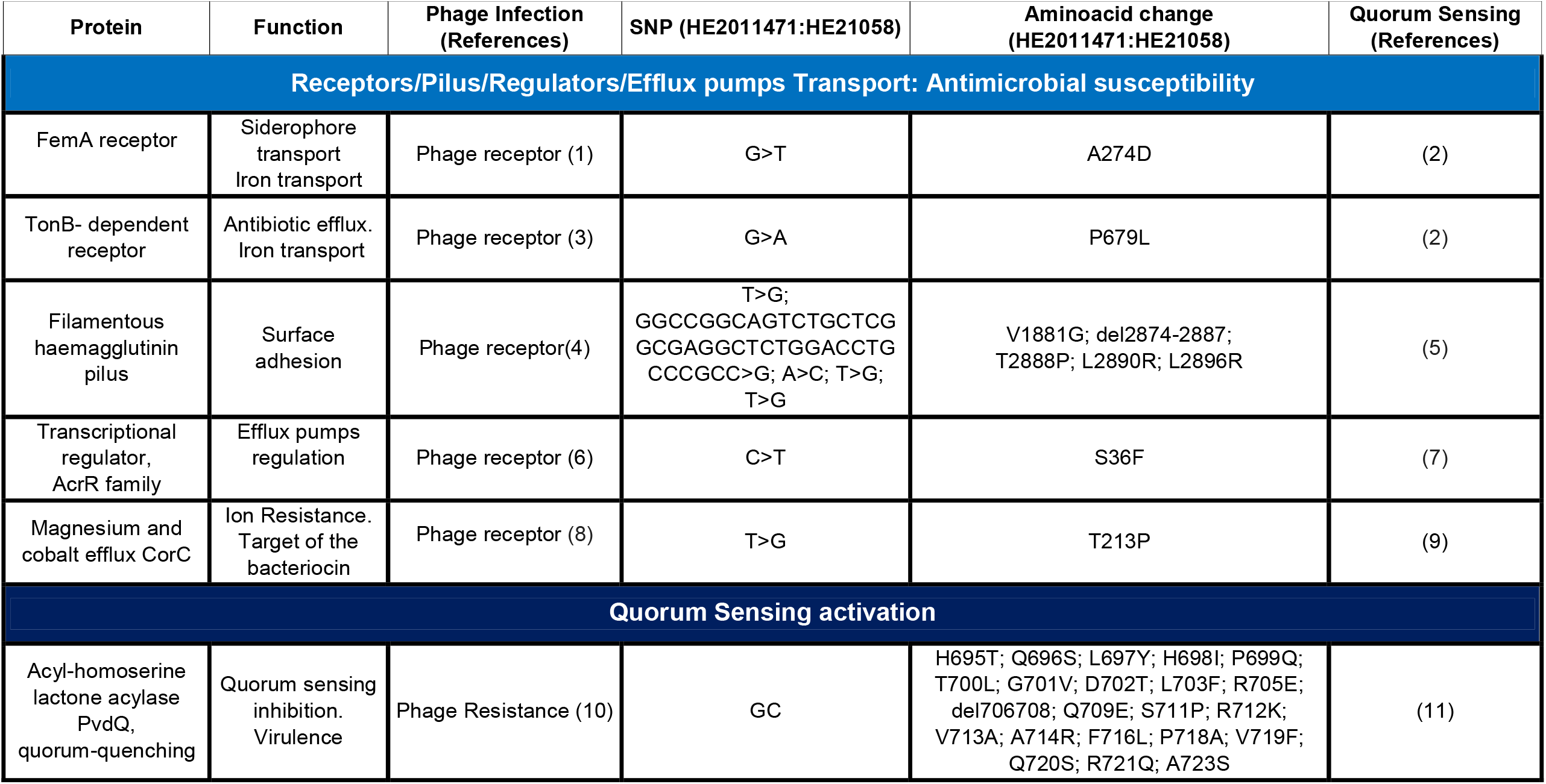
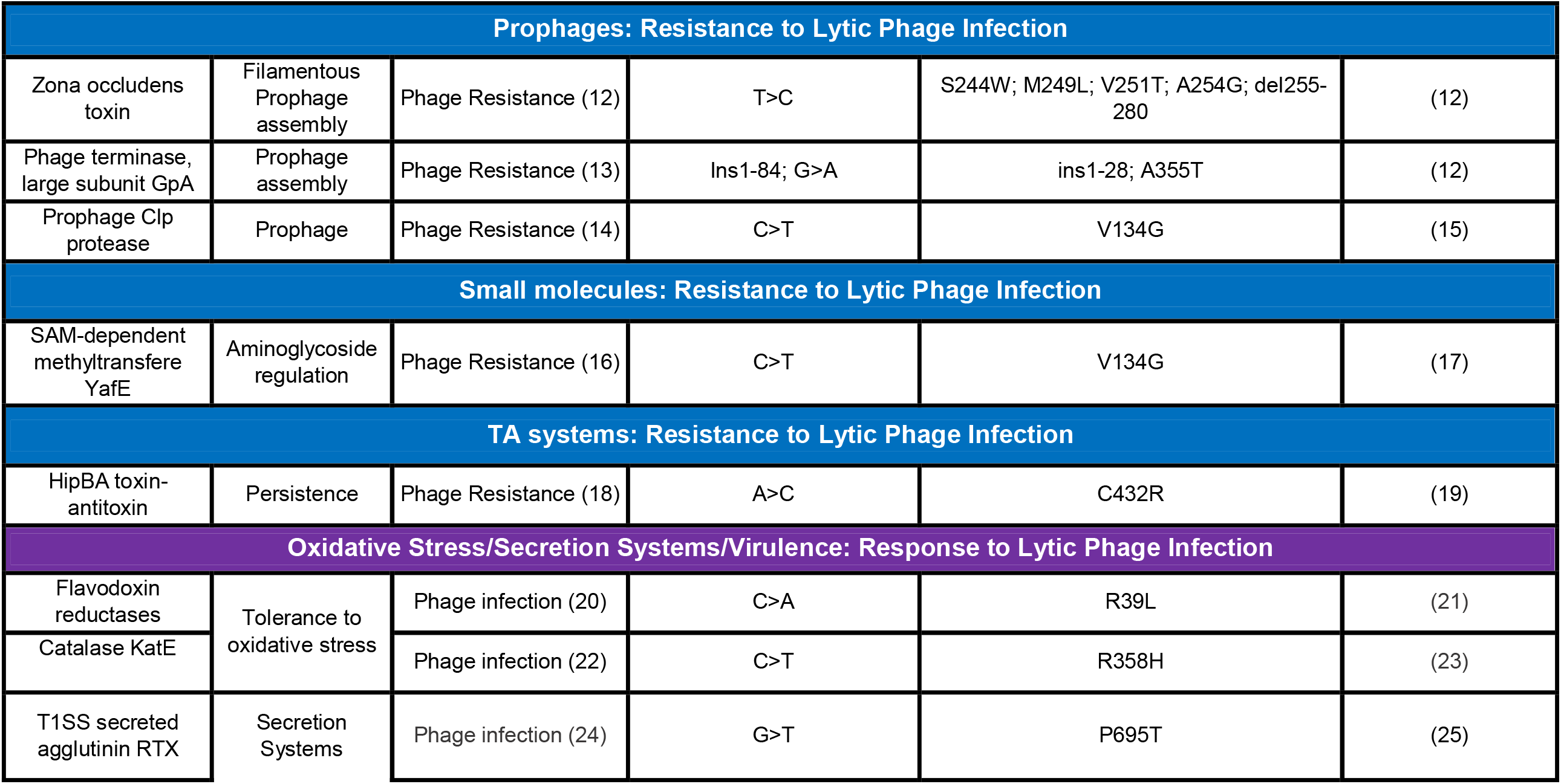
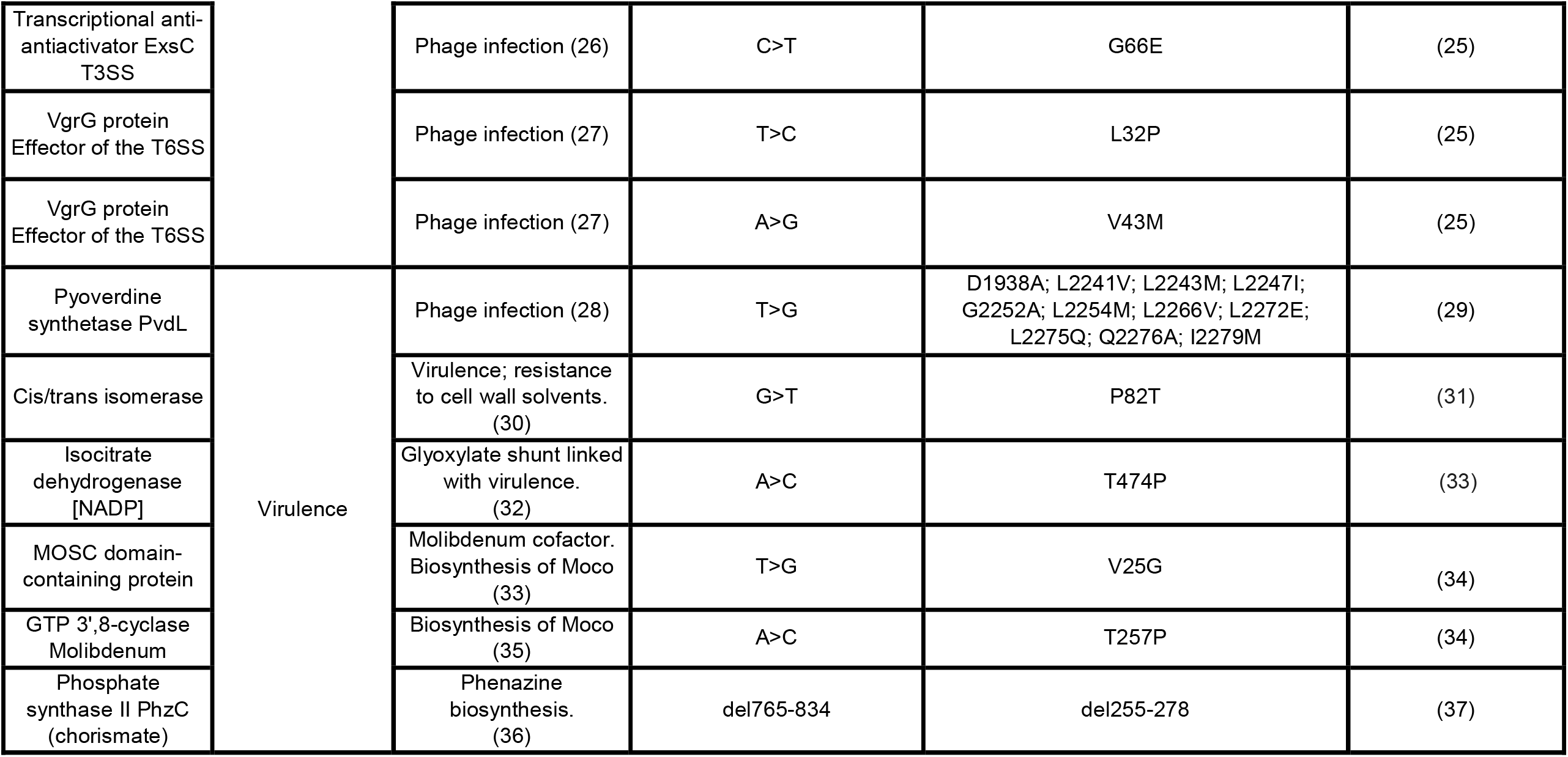
Mutations encountered in proteins supposed to be related to phage resistance in response to lytic phage infection in *P. aeruginosa*. ^a^ Proteins located in prophages as identified by Phaster and SourceFinder.

